# Automated Detection of Macro-Reentrant Atrial Tachycardia Circuits Using LAT-Derived Graph Networks

**DOI:** 10.64898/2026.04.01.26350012

**Authors:** Marcus Talke, Jonah Majumder, Michael Lavelle, Sarah Schwartz, Edward J. Ciaccio, Hirad Yarmohammadi, Geoffrey Rubin, Jessica A. Hennessey, Angelo B. Biviano, Hasan Garan, Elaine Y. Wan, Seth Goldbarg, Joon Hyuk Kim, Christine Hendon, Deepak Saluja

## Abstract

**Background:** Accurate identification of macro-reentrant atrial tachycardia (AT) circuits is critical for successful ablation but remains challenging with conventional mapping techniques. The aim of this study was to automatically detect macro-reentrant AT loops from high-density local activation time (LAT) maps.

**Methods:** We developed an algorithm for automated detection of macro-reentrant AT circuits using LAT-derived directed graphs. Compared to previous graph-based approaches, the algorithm is designed to identify the fastest-conducting reentrant pathways and cluster them by rotational orientation (clockwise vs. counterclockwise) to distinguish single- from dual-loop circuits. The algorithm was applied retrospectively to 60 macro-reentrant scar-related AT cases mapped with CARTO or Ensite from two institutions. The results were compared with blinded expert electrophysiologist annotations of loop location and single- vs. dual-loop classification.

**Results:** The 60 cases included 16 right atrial and 44 left atrial ATs from 51 patients. Expert review identified 57% single-loop and 43% dual-loop circuits. Compared with expert annotation, the algorithm correctly identified anatomical loop locations with 88% accuracy and correctly distinguished single- vs. dual-loop ATs in 93% of cases.

**Conclusion:** Our LAT graph-based algorithm automatically identified single- and dual-loop macro-reentrant AT circuits. Localizing these pathways may provide insight into circuit mechanisms and help guide ablation.

**CLINICAL PERSPECTIVE:** **What is Known:**

- Accurate characterization of macro-reentrant atrial tachycardias (ATs) is essential for effective catheter ablation, yet identifying these circuits from local activation time (LAT) maps remains challenging and operator dependent.

**What the Study Adds:**

- We developed and validated a graph-based algorithm that automatically identifies macro-reentrant AT circuits from LAT maps with high agreement compared to expert electrophysiologist annotation.
- The algorithm localizes reentrant loop pathways and distinguishes single- from dual-loop circuits.
- Localizing these pathways may provide insight into circuit mechanisms and help guide ablation.

## INTRODUCTION

Macro-reentrant atrial tachycardias (ATs) are organized electrical circuits circulating around anatomical or functional barriers such as valve annuli, surgical scars, or areas of fibrosis [1,2]. Effective catheter ablation relies on accurate visualization and mechanistic understanding of these reentry loops. High-density local activation time (LAT) mapping provides detailed spatiotemporal representations of atrial activation. However, interpretation of these maps remains largely qualitative and operator dependent. Thus, automated reentry circuit detection and characterization are desirable to reduce operator variability.

Several computational approaches have been developed from LAT maps. For example, vector field visualizations illustrate general activation flow but still require visual interpretation to infer circuit pathways [3,4]. Other tools have focused on localizing slow conduction zones as potential isthmus sites; however, these approaches lack the global context of the entire circuit, making it difficult to distinguish the critical isthmus from bystander regions of slow conduction [5,6,7,8,9].

Recently, directed graph mapping (DGM) has been proposed to automatically detect macro-reentrant loops by representing cardiac activation as a directed network derived from spatially uniform sampled LAT maps [10,11]. This approach demonstrates the promising potential of graph-based analysis for objective identification of reentry circuits.

In addition, recent studies have revealed that macro-reentrant ATs often exhibit dual-loop configurations sharing a common isthmus [12,13,14]. Characterizing loop rotational orientation may facilitate recognition of dual-loop circuits and their shared pathways that sustain arrhythmia propagation. While some graph-based methods such as DGM-TOP can compute rotational direction, they still require manual annotation of anatomical or functional rotational boundaries [15].

To address some of these challenges, we developed a novel graph-based algorithm for automated detection of macro-reentrant pathways from LAT maps. Our approach incorporates three key methodological innovations: (1) *spatiotemporal sampling via wavefront tracking* to best capture activation dynamics, enabling (2) *physiologically informed path selection*, which identifies the fastest conducting pathways under the hypothesis that the dominant reentrant circuit corresponds to the fastest propagation route that sustains the tachycardia, and (3) *rotational clustering*, which groups detected loops by clockwise (CW) versus counterclockwise (CCW) orientation, facilitating identification of dual-loop circuits.

The aim of this study is to develop and validate an automated method for detecting macro-reentrant circuits in ATs, focusing on accurate loop localization and characterization of single-versus dual-loop circuit patterns.

## METHODS

### Data Acquisition

Data were collected from 62 electrophysiological ablation cases from 53 patients between February 2021 and February 2023. The study was approved by the Columbia University Irving Medical Center Institutional Review Board (IRB-AAAT8818). We included data from patients who were 18 years or older with scar-related, single-chamber macro-reentrant AT who underwent catheter ablation in an electrophysiological lab from two different institutions. Each center’s authors attested to the integrity of their institutional data.

Procedures were performed under general anesthesia or moderate sedation by one of three electrophysiologists (DS, HY, AB). Electroanatomic mapping was performed using either the CARTO 3 system with multi-spline catheters (Pentaray; 20-poles with 3 mm spacing or Octaray; 48-poles with 2 or 3 mm spacing, Biosense Webster) or the Ensite system with a grid-shaped 16-pole catheter with 4 mm spacing (HD Grid; Abbott Cardiovascular). Ablation was performed with either the SmartTouch Surround Flow (Biosense Webster) or TactiCath (Abbott Cardiovascular) irrigated radiofrequency catheters, or with the FARAPULSE (Boston Scientific) pulsed field ablation catheter. Arrhythmias were terminated with ablation in all patients, and ablation sites were recorded.

For each case, LATs, voltage, and Cartesian coordinates (x,y,z) of electrode sites were recorded during mapping. LATs were automatically annotated by the electroanatomic mapping systems. LATs were determined using maximum dV/dT for Ensite Precision, peak frequency for Ensite X, and a proprietary algorithm based on maximal –dV/dT for CARTO 3. Data was exported from the electroanatomic mapping system for offline analysis.

### Algorithm

We developed a graph-based algorithm to identify macro-reentrant circuits from high-density LAT maps. The algorithm detects looped paths corresponding to the fastest conducting pathways within the circuit, motivated by the hypothesis that, among all possible reentrant pathways, the physiologically dominant circuit corresponds to the fastest propagation route sustaining the tachycardia. These paths avoid conduction block, which is characterized by steep LAT gradients, but traverse slow-conduction zones, such as the isthmus, when necessary to complete the circuit. Below is a summary of the algorithm with full technical details provided in the Appendix.

#### 1. Input data and preprocessing

The algorithm inputs were LATs (ms) at 3D Cartesian coordinates (x,y,z), the tachycardia cycle length, and an approximately equilateral triangulated surface mesh. LAT values were normalized to the tachycardia cycle length and converted into circular phase (0–360°). Phases were projected and interpolated on the anatomical mesh to create a continuous activation map.

#### 2. Graph Construction

Normalized LAT phases were discretized into 24 bins of 15° each. Mesh vertices that were anatomically connected and belonged to the same phase bin were grouped into subgraphs, referred to as *islands*, which represent discretized wavefronts of electrical propagation. Each island was summarized by a single node at its centroid, and directed edges between islands were assigned along increasing phase, representing anterograde conduction. These steps constitute a form of spatiotemporal sampling, exemplified schematically in Figure 1A-C. Edge weights were defined based on phase differences and spatial distances between islands.

**Figure 1.**
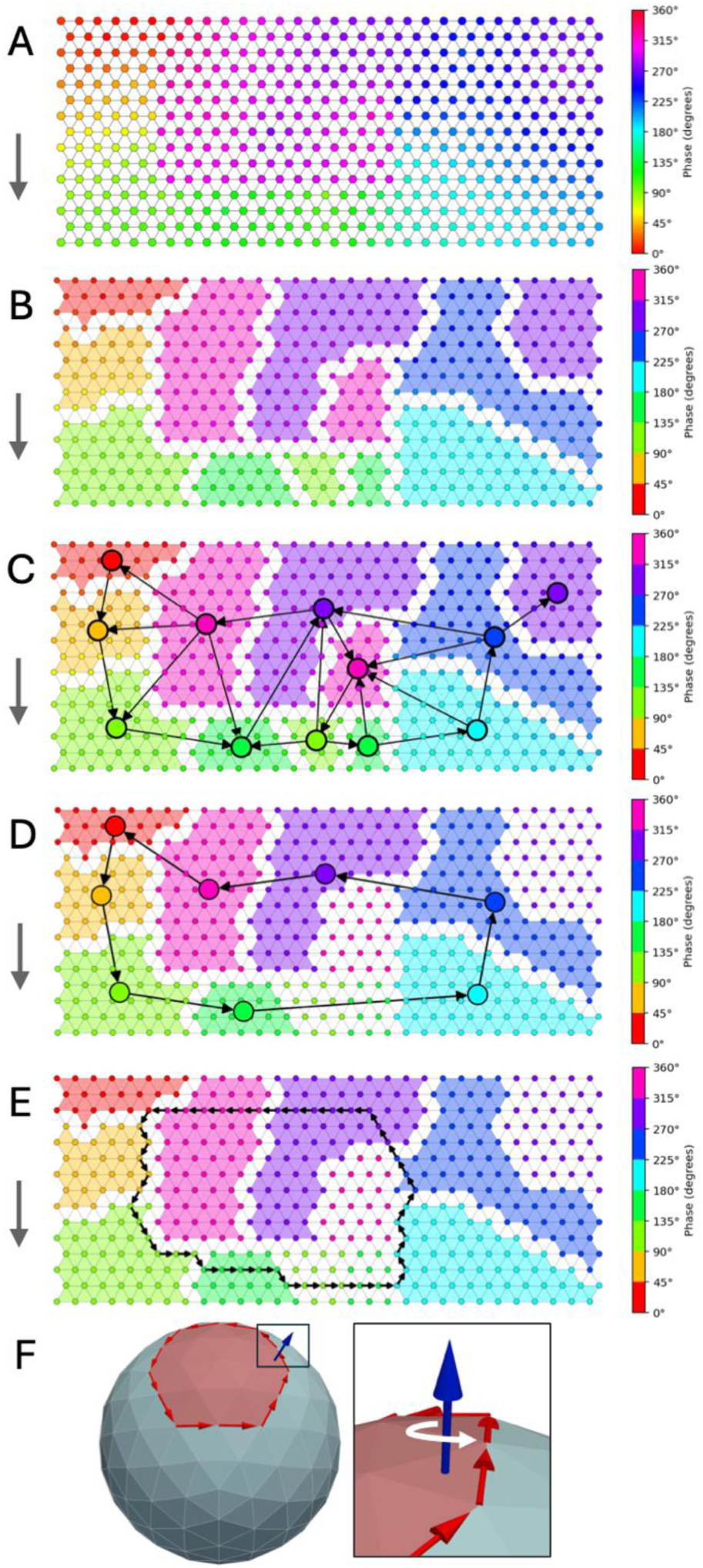
Schematic pipeline of algorithm. **A,** triangulated mesh with interpolated LAT (converted to 0-360 degree phase). **B,** isochronal binning to form islands. **C,** island graph construction with node centered on its respective island subgraph. **D,** algorithm finds island loop that is fastest-conducting pathway. **E,** island loop is mapped back to original anatomical mesh to find the shortest pathway. **F,** the loop interior (smaller enclosed surface area in red) was first identified, and rotational orientation was then computed relative to the local interior surface normal vector in blue to classify rotational orientation (CW vs. CCW).

#### 3. Loop Detection

A Dijkstra-inspired lexicographic best-first search algorithm was developed to identify island loops with the fastest conduction speed (Figure 1D). These island loops were then mapped onto the anatomical mesh, producing a continuous path along mesh vertices corresponding to the shortest reentrant circuit (Figure 1E).

Unlike classical Dijkstra’s algorithm, which minimizes a scalar additive cost along a path, we used lexicographic comparison of vector-valued path weights. This framework prioritizes avoiding large phase jumps across block, whereas Dijkstra’s algorithm may accept high-cost transitions across block if compensated elsewhere along the path.

#### 4. Loop Classification and Selection

Detected loops were then clustered by rotational orientation (CW vs. CCW) with respect to the loop interior (smallest enclosed loop area). Rotational orientation was determined by comparing the cross product of the loop vector and an inward-pointing vector with the local surface normal; alignment indicated CCW orientation, and opposition indicated CW orientation (Figure 1F). The loop with the fastest conduction speed was selected in each cluster, with the shortest loop used as a tiebreaker, yielding at most one CW and one CCW loop.

### Algorithm Validation

Algorithm performance was evaluated with respect to blinded expert review performed offline. Two electrophysiologists (DS, ML) independently and blindly reviewed each case to localize the macro-reentrant AT circuit and determine whether the arrhythmia mechanism was single- or dual-loop using LAT maps, voltage maps, conduction speed maps, and ablation data. When circuit localization was ambiguous, reviewers prioritized loop pathways consistent with ablation termination sites when present. Discrepancies were resolved by consensus. Cases without consensus were excluded.

Macro-reentrant loops were defined as circuits rotating around a central obstacle >2 cm in diameter [16,17]. In this study, all ATs were scar-mediated. However, for descriptive purposes, loops were categorized according to the anatomical structure they encircled, providing a simplified reference for circuit localization. In the RA, loops were classified as encircling the superior vena cava (SVC), inferior vena cava (IVC), tricuspid valve (TV), or scar tissue not involving an anatomical landmark. In the LA, loops were classified as encircling the left pulmonary veins (LPVs), right pulmonary veins (RPVs), mitral valve (MV), or scar tissue not involving an anatomical landmark. Figure 2 illustrates these loop categories schematically in a cartoon representation of the atria.

**Figure 2.**
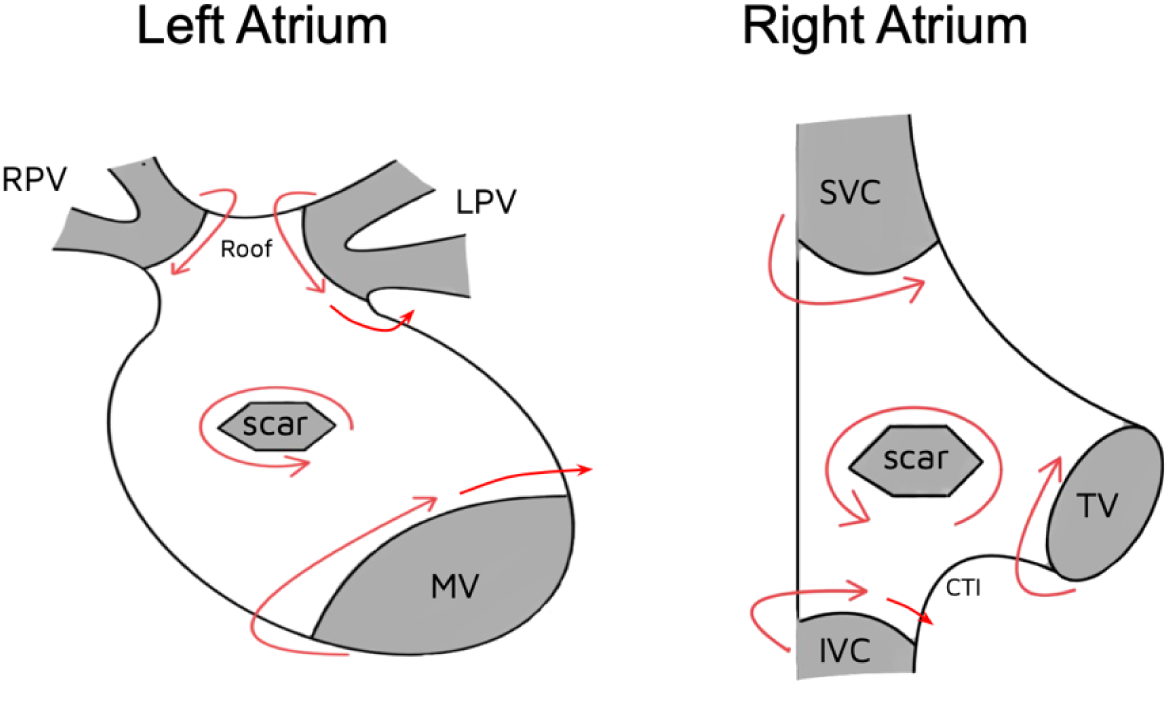
Classification of macro-reentrant AT loops by anatomic landmarks. The LA circuit locations are LPV, RPV, MV, and scar tissue. The RA circuit locations are SVC, IVC, TV, and scar tissue. Note that in literature, IVC and TV loops are commonly known as CTI-dependent and LPV and RPV loops are commonly known as roof-dependent.

Algorithm performance was evaluated using two metrics: the mean per-case Jaccard index quantifying agreement between algorithm-identified and expert-defined loop locations, and the percentage of cases correctly classified as single- or dual-loop. The Jaccard index was computed as |*A* ∩ *B*|/|*A* ∪ *B*|, where A and B represent the sets of loop locations (e.g., MV, LPV) identified by the algorithm and experts, respectively. For example, if both identified a MV loop only, the index equals 100% (perfect agreement); if the expert identified a MV loop and the algorithm identified both MV and LPV loops, the index equals 50% (one shared location out of two total).

The algorithm’s total runtime was measured per case and also partitioned into key computational steps: island loop computation (Figure 1B–D), anatomical mesh loop construction (Figure 1E), and rotational clustering (Figure 1F).

### Statistical Analysis

Continuous variables are reported as mean ± standard deviation (SD) or median with interquartile range (IQR), depending on data distribution. Categorical variables are expressed as counts and percentages. For the per-case Jaccard index, 95% confidence intervals (CIs) were estimated using bootstrap resampling with 10,000 iterations. All analyses were performed using Python (version 3.13; Python Software Foundation, Wilmington, DE) with SciPy.

## RESULTS

### Cohort and Case Overview

Of 62 AT cases from 53 patients, two cases (two patients) were excluded because the expert electrophysiologists could not reach consensus. The final cohort comprised 60 AT cases from 51 patients, including 16 right atrium (RA) and 44 left atrium (LA) tachycardias. The demographic data of the subjects are listed in Table 1. The median number of LAT points per case was 3,665 [IQR 2,024–5,280], and the median cycle length of ATs was 270 ms [IQR 236–303]. Of these cases, 6 (10%) were mapped with Ensite Precision, 25 (41.7%) with Ensite X, and 29 (48.3%) with CARTO.

**Table 1.** Demographic characteristics of the 51 patients.

| **Characteristic** | **Count (%)** |
| --- | --- |
| **Age (yrs)** |  |
| <50 years | 4 (7.8%) |
| 50-59 years | 5 (9.8%) |
| 60-69 years | 19 (37.3%) |
| 70-79 years | 16 (31.4%) |
| >= 80 years | 7 (13.7%) |
| **Sex** |  |
| Male | 34 (66.7%) |
| Female | 17 (33.3%) |
| **Race** |  |
| White | 29 (56.9%) |
| Black | 5 (9.8%) |
| Asian | 2 (3.9%) |
| Other | 9 (17.6%) |
| Unspecified | 6 (11.8%) |
| **Body Mass Index** |  |
| < 18.5 | 1 (1.9%) |
| 18.5-24.9 | 11 (21.6%) |
| 25-29.9 | 22 (43.1%) |
| >= 30 | 17 (33.3%) |
| **Comorbidities** |  |
| Hypertension | 46 (90.2%) |
| Hyperlipidemia | 42 (82.4%) |
| Congestive heart failure | 25 (49%) |
| Coronary artery disease | 24 (47.1%) |
| Diabetes mellitus | 12 (23.5%) |
| Stroke | 7 (13.7%) |

### Blinded Expert Review

Expert review identified 57% single-loop (34/60) and 43% dual-loop (26/60) circuits. In the RA (n = 16), loops involved the SVC in 3 cases (18.8%), the IVC in 3 cases (18.8%), the TV in 3 cases (18.8%), and scar tissue alone in 11 cases (68.8%). In the LA (n = 44), loops involved the MV in 27 cases (61.4%), the LPV in 16 cases (36.4%), the RPV in 11 cases (25.0%), and scar tissue alone in 11 cases (25.0%). Figure 3 shows overall concordance between expert-reviewed and algorithm-predicted loop classifications, illustrated on a case-by-case basis. Misclassifications primarily arose when differentiating scar-only loops from loops bounded by anatomical structures that included scar, particularly in the RA. These discrepancies were largely attributable to differences in LAT gradient interpretation between the algorithm and expert annotations. Additional discrepancies involved single loops labeled as dual or, rarely, dual loops labeled as single, with the algorithm always capturing at least one expert-identified loop correctly.

**Figure 3.**
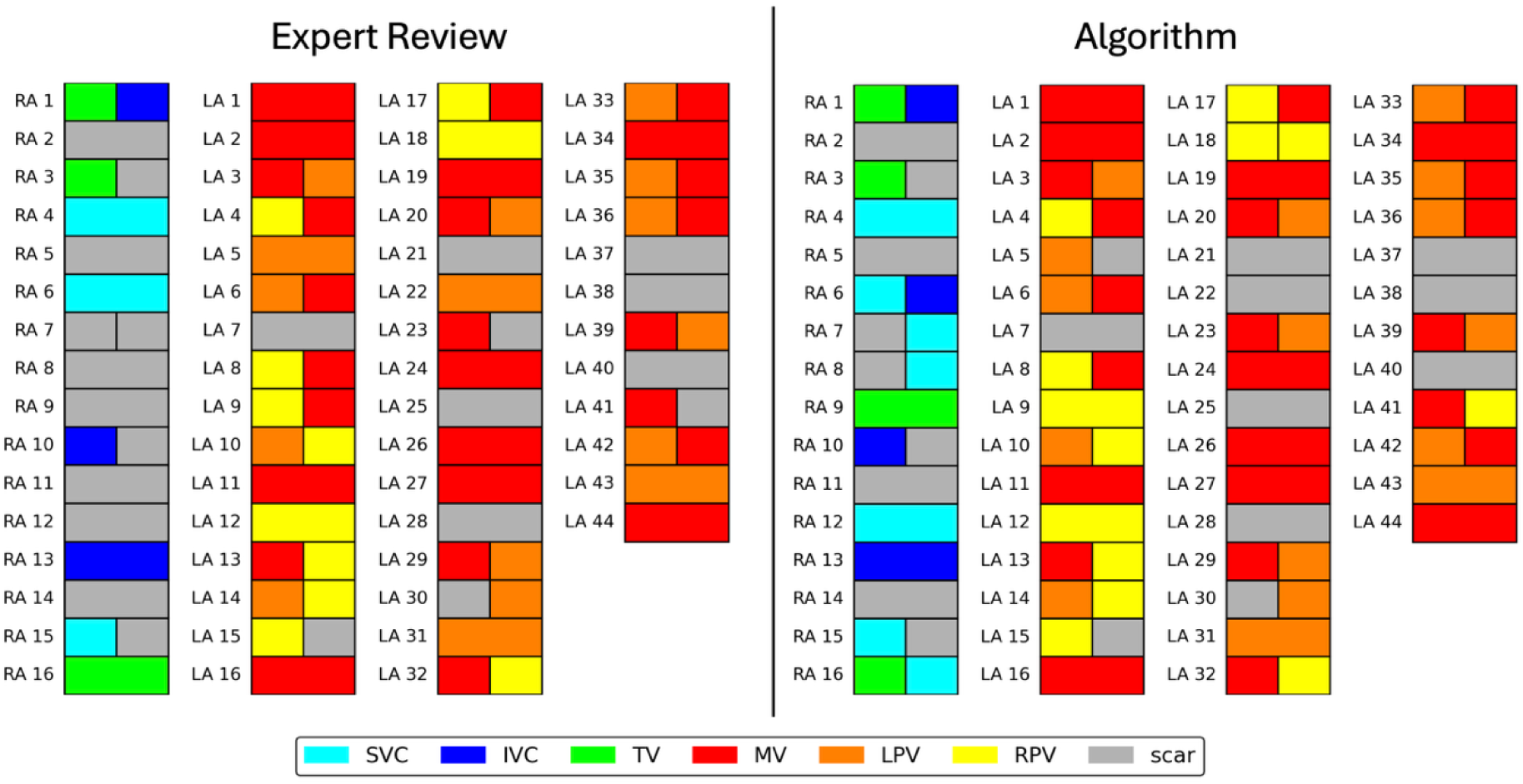
Per-case comparison of expert and algorithm loop classification. Side-by-side comparison of macro-reentrant loop classification by expert review (left) and the algorithm (right). Each row represents a single case. The color of the cells denote the anatomical structure encircled by the identified loop (SVC, IVC, TV, MV, LPV, RPV, or scar). Split cells indicate dual-loop circuits.

### Algorithm Performance

The algorithm achieved a mean per-case Jaccard index of 87.8% (95% CI: 80.3–94.2%) for loop localization and correctly identified a single- vs. dual-loop mechanism in 93.3% of cases when compared to expert review. Figure 4 compares expert-reviewed and algorithm-predicted loop classifications by anatomical location and single- versus dual-loop mechanism. Overall concordance was high. Misclassification was more frequent in the RA than LA. The algorithm tended to overidentify dual-loop cases and undercount single-loop cases.

**Figure 4.**
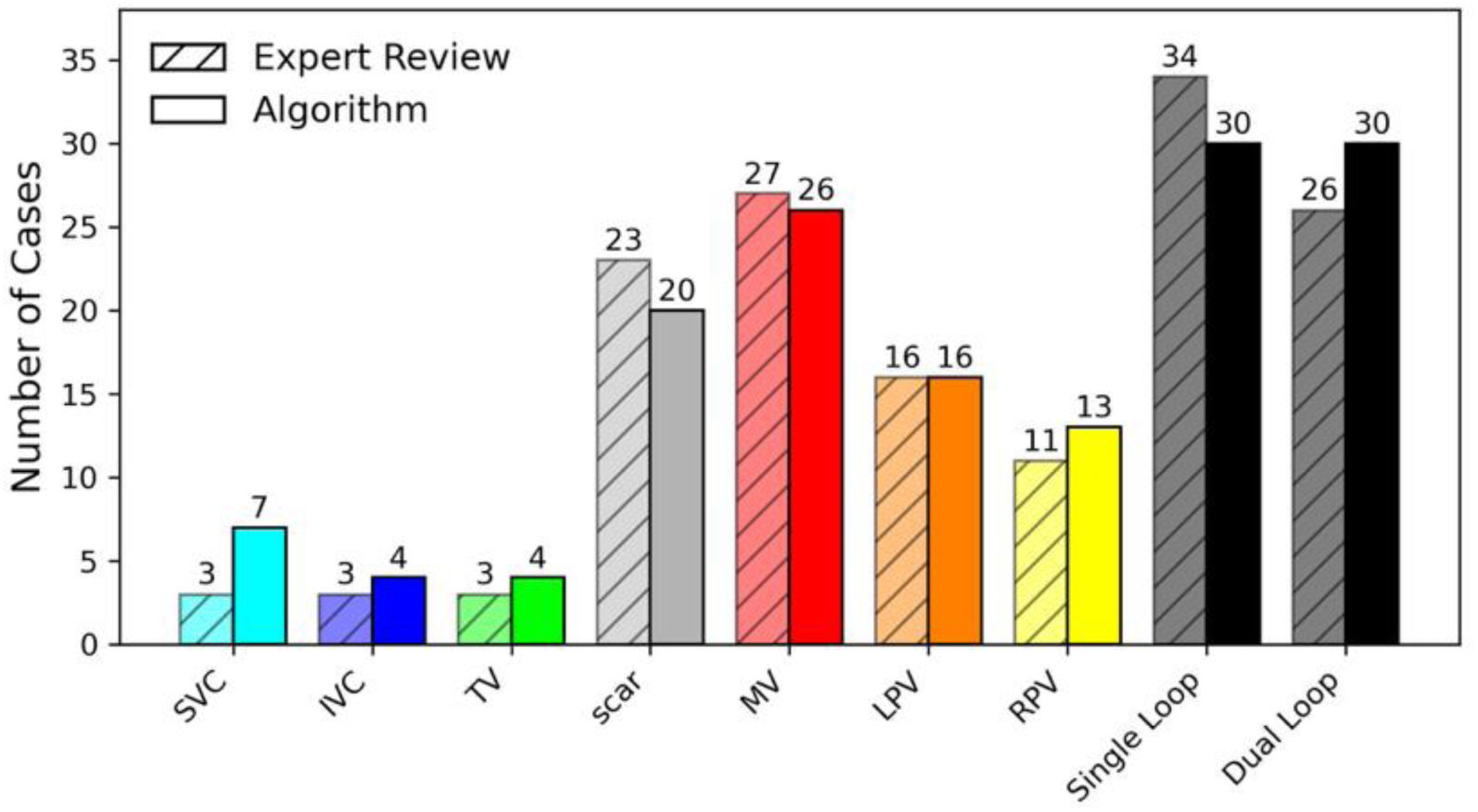
Expert review vs. algorithm: loop classification by anatomical location. Comparison of expert-reviewed and algorithm-predicted loop classifications. Bars show the number of cases identified for each anatomical location and for single- vs. dual-loop mechanism.

The algorithm’s median total runtime was 6.78 s (IQR 4.08–9.08 s). Stepwise analysis showed median runtimes of 0.31 s (IQR 0.13–0.70 s) for computing island loops, 2.59 s (IQR 1.53–3.48 s) for anatomical mesh loops, and 3.83 s (IQR 2.41–5.17 s) for rotational clustering.

### Examples of Algorithm-Identified Single-Loop Circuits

Figure 5 illustrates a RA case in which the algorithm identified a single-loop circuit rotating around scar tissue in the posterior wall. The algorithm-derived CCW loop is superimposed on the LAT map (left), on a LAT map highlighting only circuit wavefronts with bystander activation darkened (middle), and on a conduction speed map (right). The conduction speed panel (right) shows the loop rotating around block while identifying the fastest-conducting breakthrough sites across lines of block to complete the circuit.

**Figure 5.**
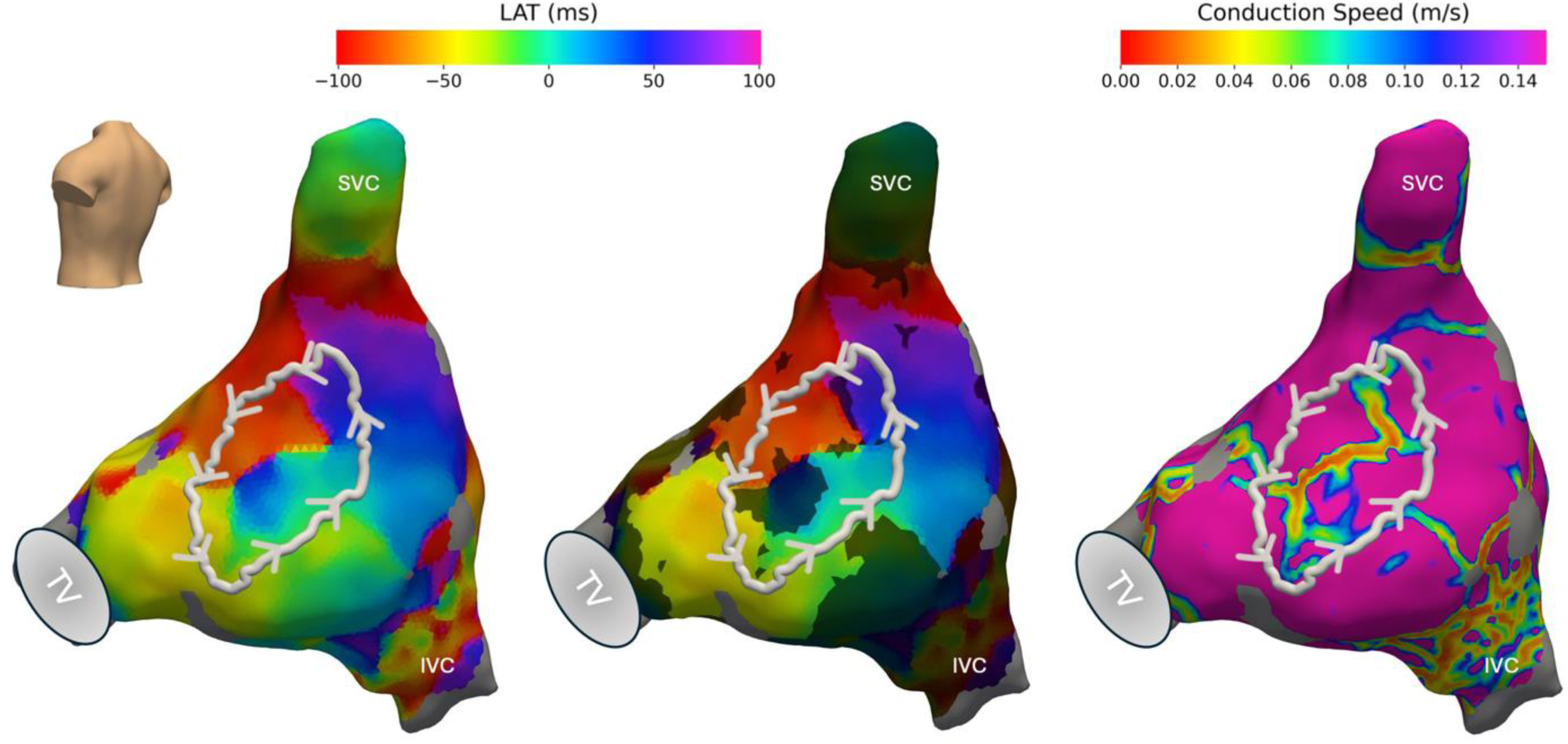
First example of single-loop. RA single-loop reentry is shown, with the white loop rotating CCW around scar on the posterior wall. Left to right: LAT map, LAT map highlighting wavefronts participating in circuit (with other regions dimmed), and conduction speed map.

Similarly, Figure 6 shows another single-loop circuit in the LA rotating around the MV. The algorithm-derived CW loop is superimposed on the LAT map (left), on a LAT map highlighting only circuit wavefronts with bystander activation darkened (middle), and on a conduction speed map (right). The middle panel highlights the bottleneck shape of the critical isthmus, which was ablated to terminate the circuit. The conduction speed panel (right) shows the algorithm localizing the fastest conducting pathway while finding breakthrough across lines of block.

**Figure 6.**
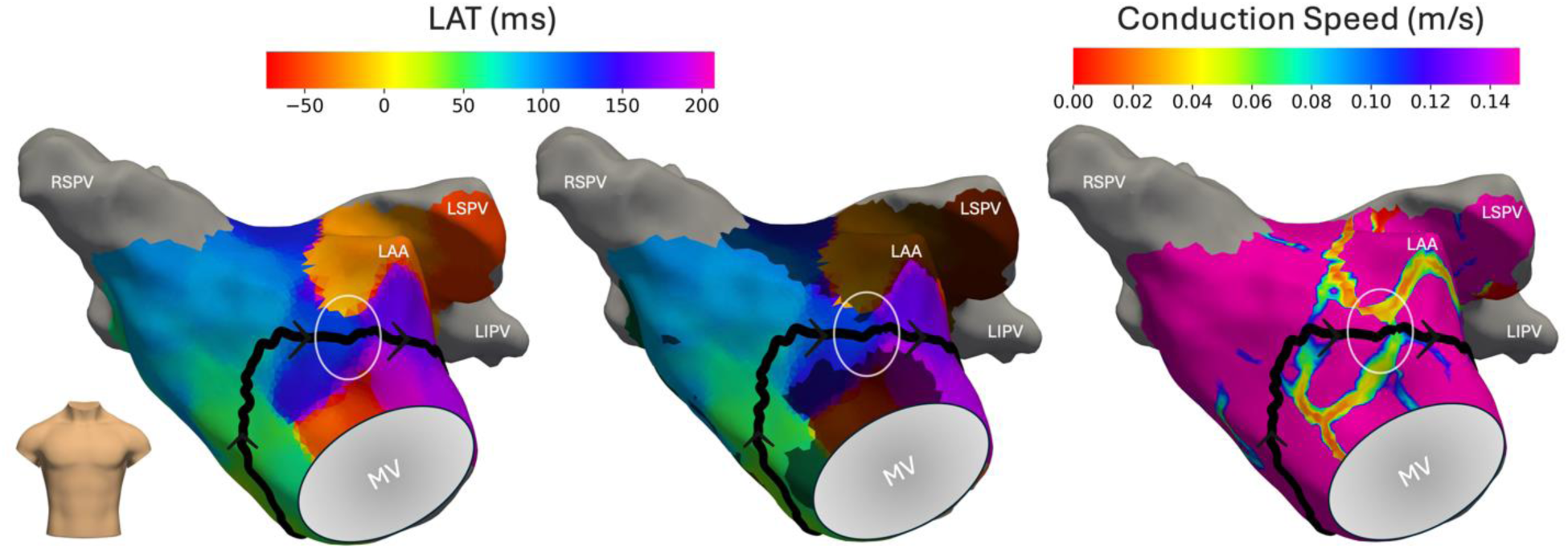
Second example of single-loop. LA single-loop reentry is shown, with the black loop rotating CW around the MV. Left to right: LAT map, LAT map highlighting wavefronts participating in circuit (with other regions dimmed), and conduction speed map. The critical isthmus, circled in white, shows a characteristic bottleneck shape, most evident in the middle panel. RSPV = right superior pulmonary vein; LSPV = left superior pulmonary vein; LIPV = left inferior pulmonary vein; LAA = left atrial appendage.

### Examples of Algorithm-Identified Dual-Loop Circuits

Figure 7 shows a dual-loop circuit in the LA, with two algorithm-derived loops superimposed on the LAT map (left), a transparent anatomical map (middle), and a conduction speed map (right). The CW (black) loop rotates around posterior scar, and the CCW (white) loop rotates around the LPVs, sharing a common pathway (striped segment) along the critical isthmus.

**Figure 7.**
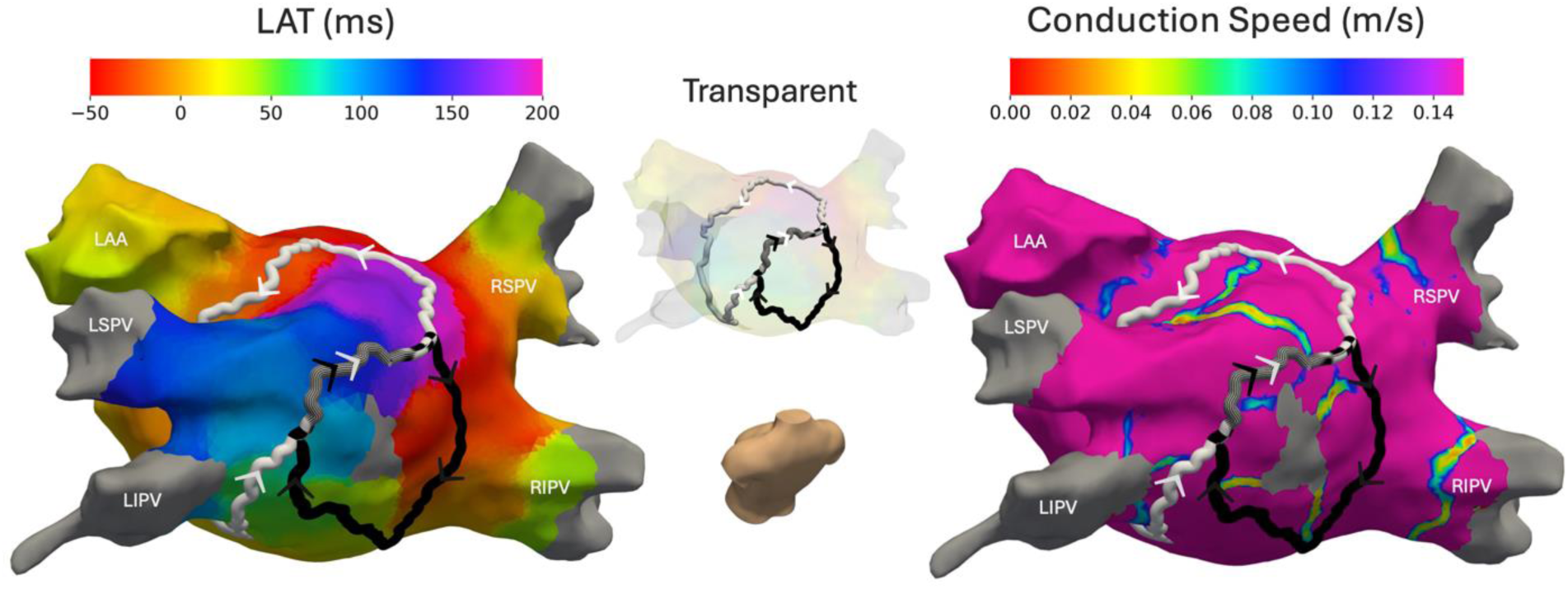
First example of dual-loop. Left to right: activation map, transparent map, and conduction speed map. LA dual-loop reentry is shown, with the white loop rotating CCW around the LPVs and the black loop rotating CW around posterior wall scar. The shared pathway (striped segment) was ablated, terminating the arrhythmia. RSPV = right superior pulmonary vein; RIPV = right inferior pulmonary vein; LSPV = left superior pulmonary vein; LIPV = left inferior pulmonary vein; LAA = left atrial appendage.

Figure 8 shows a dual-loop circuit in the LA, with an algorithm-identified CCW loop (white) around the MV and a CW loop (black) around the RPVs. These loops are shown on the LAT map (left), conduction speed map (middle), and transparent map (right). However, expert review disagreed with the algorithm and identified a shorter CW loop around scar alone (dotted arrow) rather than extending around the RPVs. Regardless, the shared pathway was ablated with successful termination of the circuit.

**Figure 8.**
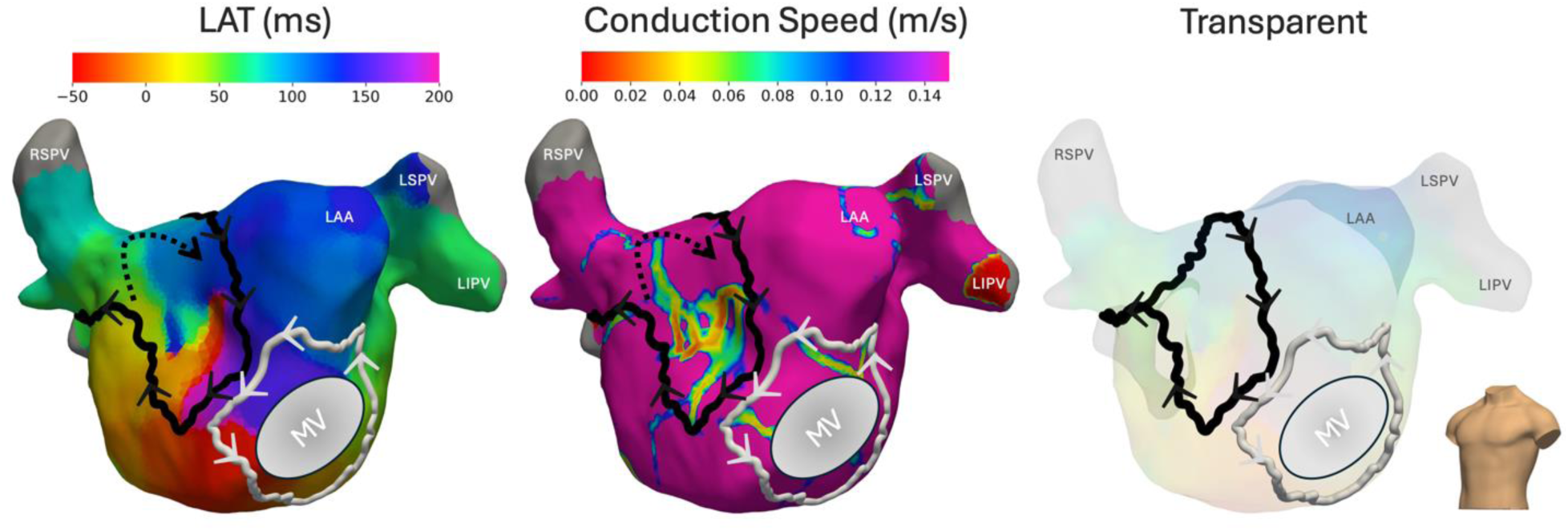
Second example of dual-loop. Left to right: LAT map, conduction speed map, and transparent map. The algorithm-predicted LA dual-loop reentry is shown, with the white loop rotating CCW around the MV and the black loop rotating CW around the right superior pulmonary vein (RSPV). Expert review identified a shorter alternative CW loop around scar only (dotted black arrow). The shared pathway was ablated, terminating the arrhythmia. LAA = left atrial appendage; LSPV = left superior pulmonary vein; LIPV = left inferior pulmonary vein.

The final example in Figure 9 shows a dual-loop circuit in the LA with a CCW loop (white) around the MV and CW loop (black) around the LPVs. The algorithm-identified loops are shown on the LAT map (left), conduction speed map (middle), and transparent map (right). The middle panel shows both loops forming a shared pathway (striped segment) finding the fastest-conducting breakthrough site across the line of block.

**Figure 9.**
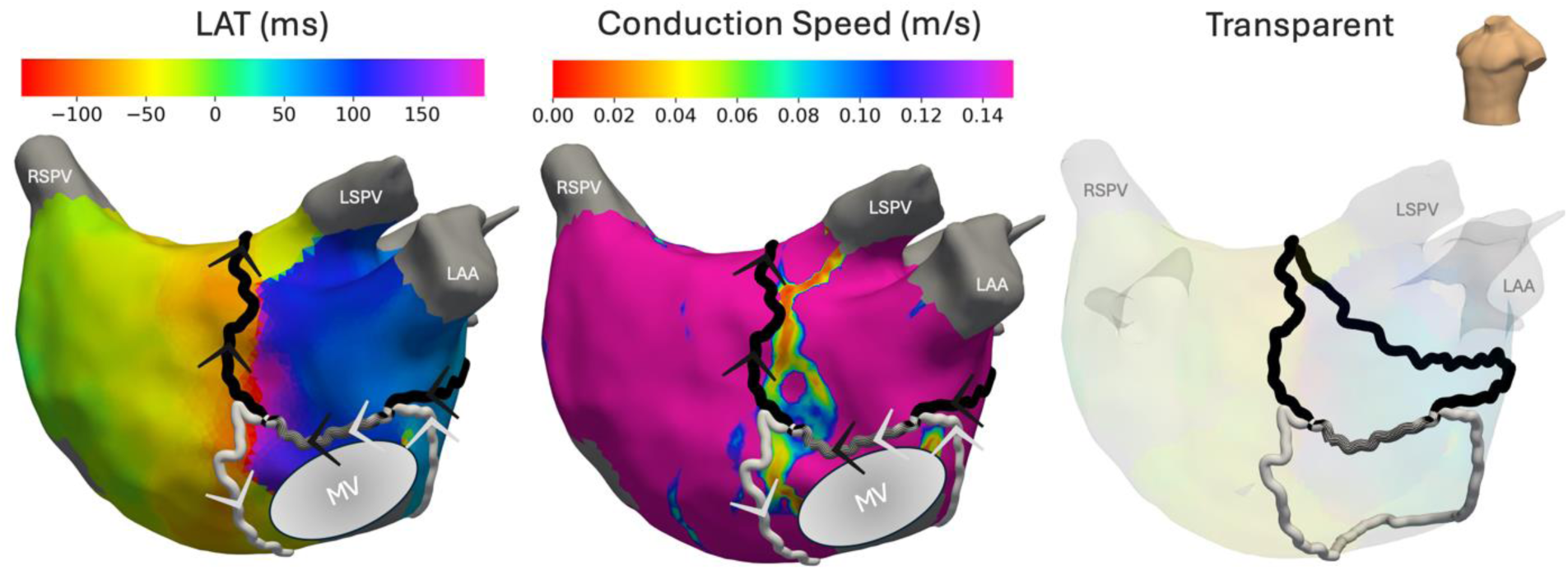
Third example of dual-loop. Left to right: activation map, conduction speed map, and transparent map. LA dual-loop reentry is shown, with the white loop rotating CCW around the MV and the black loop rotating CW around the LPVs. LAA = left atrial appendage; RSPV = right superior pulmonary vein; LSPV = left superior pulmonary vein.

## DISCUSSION

In this study, we present an automated graph-based algorithm for detecting and characterizing macro-reentrant AT circuits from high-density LAT maps. By finding the fastest conducting pathways and clustering by rotational orientation, the algorithm localized reentrant pathways with 87.8% accuracy and distinguished single- from dual-loop mechanisms with 93.3% accuracy when compared to blinded expert review. These findings suggest that this algorithm can reproducibly localize and characterize macro-reentrant circuits, potentially reducing operator variability and improving mechanistic interpretation of complex ATs.

Other investigators have shown promising results using graph-based methods for localizing macro-reentrant circuits [10,11]. However, several key challenges remain. DGM relies on equal spatial sampling, which may overlook complex temporal wavefront dynamics; it minimizes LAT step variance, a criterion with an unclear physiologic basis; it does not strictly enforce surface contiguity, allowing non-anatomic pathway jumps; and it clusters loops based on distance between loop centroids, which may fail to identify spatially adjacent dual-loop circuits. Our algorithm addresses these challenges by introducing several novel features. First, we implement spatiotemporal sampling to track electrical wavefronts, which allows for more focused attention on deceleration zones, lines of block, and breakthrough. Our algorithm is physiologically inspired and hypothesis-driven, preferentially selecting the fastest-conducting reentrant pathways, which are posited to represent the dominant propagation route sustaining the arrhythmia. Unlike previous methods [10,11], we do not rely on universal strict thresholds for conduction speed to define blocks, where thresholds can vary across cases and within cases.

Recent investigations into atrial reentry topology have shown that macro-reentrant ATs often exhibit dual-loop configurations sharing a common isthmus [12,13]. To detect these dual-loop mechanisms, we cluster loops by rotational orientation rather than spatial separation, recognizing that dual-loop circuits occur as clockwise–counterclockwise pairs. While some graph-based approaches, such as DGM-TOP, can also identify rotational orientation, they still require manual annotation of rotational boundaries [15]. In addition, our algorithm enforces path contiguity on the cardiac surface while having the ability to skip over regions affected by noise, under-interpolation, or possibly even epicardial jumps, within a user-defined radius (5 cm for this study).

### Clinical Implication

This work was originally motivated by the goal of localizing the isthmus of reentrant circuits to guide ablation strategies. Our algorithm addresses this critical first step. By identifying reentrant circuits, we can distinguish circuit driver regions from bystander activation and thereby narrow the search for the isthmus as illustrated in Figure 6; the middle panel demonstrates how highlighting wavefronts allows identification of a narrow bottleneck region of slow conduction corresponding to the isthmus. The conduction speed map (right panel) shows the circuit loop finding breakthrough sites across lines of block.

In addition, our algorithm can help identify dual-loop mechanisms and shared pathways, which are often underrecognized. One study reported that for dual-loop circuits, there were higher termination rates when the common isthmus was targeted for ablation [14]. Moreover, our algorithm may help identify dual-circuit rhythms at risk of transformation to a second arrhythmia when only one limb is terminated. The algorithm has potential for real-time use, with a median total runtime of 6.78 seconds with room for improvement using parallel computation.

The most common discrepancies between the algorithm and expert review arose from interpreting steep LAT gradients to distinguish true conduction block from slow conduction. This also affected differences in coarse anatomical loop categorization as in Figure 7, in which the algorithm identified a CW loop encircling the RPVs (including posterior scar), whereas expert review localized a slightly shorter CW loop around only posterior scar (not including the RPVs). In both interpretations, the same shared pathway was identified, which was targeted for successful ablation.

### Limitations

There are several limitations in this study. The algorithm relies solely on LAT recordings, which can be affected by noisy electrograms, LAT annotation variability, and interpolation artifacts. The algorithm does not currently incorporate additional electrogram features that could improve robustness such as voltage. Algorithm performance is dependent on accurate cycle length estimation and near-complete mapping of the macro-reentrant circuits. Validation was based on expert review, which is inherently subjective. Circuit localization was crudely evaluated using discrete anatomical classification. Lastly, the algorithm is designed to detect a maximum of two concurrent loops and does not account for quadruple-loop circuits [12] or incomplete circuits.

### Future Direction

Future work should include prospective studies to evaluate the algorithm’s clinical impact. If successful, this algorithm could be extended toward automated identification of critical isthmuses for ablation guidance. Additionally, we plan to construct dual-chamber meshes to expand the approach to biatrial tachycardias and apply this method to macro-reentrant ventricular tachycardias. While the algorithm currently detects complete circuits, further work should be done to identify incomplete circuits to improve understanding of circuit transformations following ablation.

### Conclusions

We developed and validated a novel LAT graph-based algorithm that automatically identifies macro-reentrant AT circuits. By localizing the fastest-conducting reentrant pathways and distinguishing CW and CCW loop orientations, the method accurately detected both single- and dual-loop mechanisms with high agreement to expert interpretation. Automated identification of these pathways may improve mechanistic understanding of macro-reentrant ATs with the potential to guide catheter ablation strategies.

## Data Availability

The data underlying this article are not publicly available due to patient privacy and institutional restrictions.

## Acknowledgements

None.

## Non-standard Abbreviations and Acronyms

AT: atrial tachycardia
CTI: cavotricuspid isthmus
CCW: counterclockwise
CW: clockwise
DGM: directed graph mapping
IVC: inferior vena cava
LA: left atrium
LAT: local activation time
LPV: left pulmonary vein
MV: mitral valve
RA: right atrium
RPV: right pulmonary vein
SVC: superior vena cava
TV: tricuspid valve

